# Virome Sequencing Identifies H5N1 Avian Influenza in Wastewater from Nine Cities

**DOI:** 10.1101/2024.05.10.24307179

**Authors:** Michael J. Tisza, Blake M. Hanson, Justin R. Clark, Li Wang, Katelyn Payne, Matthew C. Ross, Kristina D. Mena, Anna Gitter, Sara J. Javornik-Cregeen, Juwan Cormier, Vasanthi Avadhanula, Austen Terwilliger, John Balliew, Fuqing Wu, Janelle Rios, Jennifer Deegan, Pedro A. Piedra, Joseph F. Petrosino, Eric Boerwinkle, Anthony W. Maresso

## Abstract

Avian influenza (serotype H5N1) is a highly pathogenic virus that emerged in domestic waterfowl in 1996. Over the past decade, zoonotic transmission to mammals, including humans, has been reported. Although human to human transmission is rare, infection has been fatal in nearly half of patients who have contracted the virus in past outbreaks. The increasing presence of the virus in domesticated animals raises substantial concerns that viral adaptation to immunologically naïve humans may result in the next flu pandemic. Wastewater-based epidemiology (WBE) to track viruses was historically used to track polio and has recently been implemented for SARS-CoV2 monitoring during the COVID-19 pandemic. Here, using an agnostic, hybrid-capture sequencing approach, we report the detection of H5N1 in wastewater in nine Texas cities, with a total catchment area population in the millions, over a two-month period from March 4^th^ to April 25^th^, 2024. Sequencing reads uniquely aligning to H5N1 covered all eight genome segments, with best alignments to clade 2.3.4.4b. Notably, 19 of 23 monitored sites had at least one detection event, and the H5N1 serotype became dominant over seasonal influenza over time. A variant analysis suggests avian or bovine origin but other potential sources, especially humans, could not be excluded. We report the value of wastewater sequencing to track avian influenza.

## Main Text and Results

Highly pathogenic avian influenza viruses are extremely virulent members of the H5 and H7 subtypes of Influenza A, cause systemic disease in avian species, and incur significant cost to agricultural production because of mass culling of infected animals.^1-3^ Adaptations in the hemagglutinin (HA) surface protein that influence cleavage by the furin-like proteases is thought to be a key driver of high virulence.^1,4,5^ Since 1955, most outbreaks have been caused by three lineages, with the so-called A/goose/Guangdong/1996 H5N1 lineage widespread in poultry and responsible for human infections.^4^ The 2.3.4.4b clade evolved within this lineage and has spread throughout the world, including North America via wild birds in December 2021.^2,4,6-10^ On March 25^th^ 2024, H5N1 2.3.4.4b was detected in Texas dairy cattle herds concomitantly with herds in Michigan and Kansas.^11^ The first human case of 2024 was detected shortly thereafter in Texas (March 28^th^, 2024)^12^ in an individual with exposure to symptomatic cattle. Detections have now been observed in 36 dairy herds across nine states.^13^ Another group recently reported influenza with an H5 hemagglutinin gene present in March and April of 2024 in high levels in two wastewater sheds known to accumulate industrial discharges of cow milk.^15^ Genomic comparisons of the human and cattle cases indicate a close genetic relatedness with a single amino acid substitution of E627K within the polymerase basic protein 2 (PB2) segment.^14^

Since May of 2022, the Texas Epidemic Public Health Institute (TEPHI) has been using hybrid-capture sequencing to test weekly wastewater samples throughout Texas^16^, detecting over 400 human and animal viruses to date, several of which (e.g. SARS-CoV-2, Influenza, and Mpox) correlate to clinical case data.^17^ Seasonal influenza serotypes H3N2 and H1N1 are routinely detected in TEPHI wastewater samples, and levels have corresponded to clinical case loads from May 2022 through the beginning of March 2024. Until that point, serotype H5N1 was never detected (0 out of 1,337 wastewater samples). However, in samples from March 4^th^ through April 25^th^ (most recent available data), H5N1 is detected in 9 of 10 cities, 19 (of 23) sites, and in 46 of 163 samples (Fig. 1B, C, Fig. S1). The agnostic nature of this methodology means that these signals were observed without any change to routine protocols. The abundance of H5N1 sequences has not correlated with influenza-related hospitalizations, which have continued to decline during the spring (Fig. 1B).

**Figure 1:**
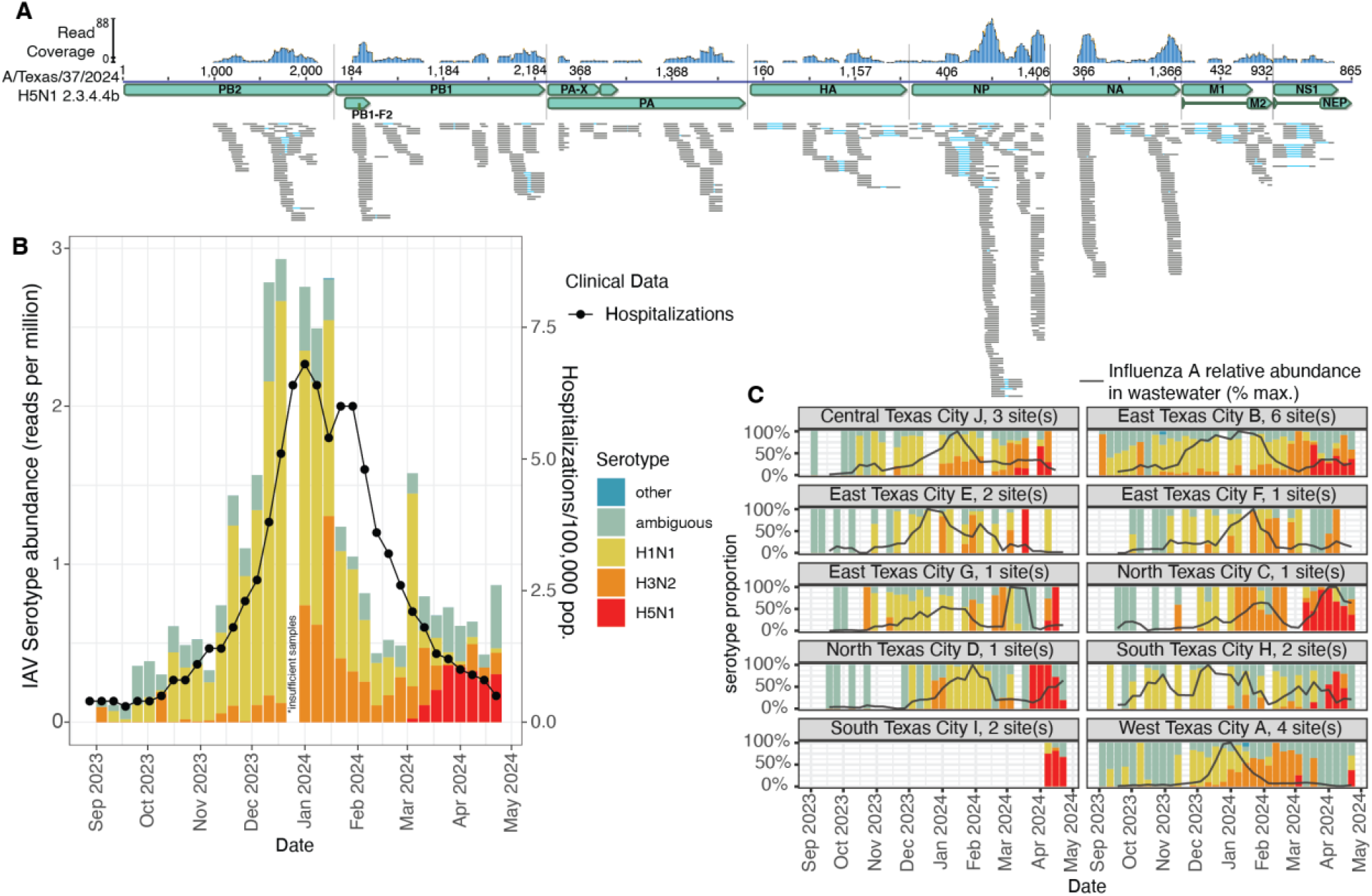
Influenza A H5N1 serotype is detectable in wastewater via virome sequencing. (A) Dereplicated reads from wastewater virome sequencing assigned to H5N1 from all samples with validated H5N1 signal (see methods) aligned to a recently collected H5N1 2.3.4.4b reference sequence (GISAID: EPI_ISL_19027114). (B) Temporal chart (September 1^st^, 2023 – April 25^th^, 2024) measuring Influenza A serotype and abundance averaged across all 23 Texas wastewater treatment sites (colored bars) and hospitalizations due to influenza across Texas as reported by Texas Department of Health and Human services (black line). Note that hospitalizations also include those caused by Influenza B, which is not measured in wastewater in this chart. (C) Bars represent proportion of influenza A serotypes in wastewater samples from each city. Grey line shows the 3-week moving average of influenza A relative abundance in wastewater, normalized to each city’s seasonal maximum. Note that sampling of South Texas City I sites began April 8^th^, 2024.

Sequences were assigned to H5N1 by a competitive alignment approach to an extensive influenza genome database, requiring read pairs to align closely to H5N1 references and better to H5N1 than other serotypes. Putative H5N1 sequences were then manually validated and documented by three independent genomics researchers associated with TEPHI (see methods). Within the set of validated H5N1 sequencing reads, there are alignments to all eight genome segments including the PB2 (on segment 1), HA/hemagglutinin (on segment 4) and NA/neuraminidase (on segment 6) genes (Fig. 1A). All sequencing reads best match H5N1 genomes from birds and mammals (including the human case) collected since 2023 and are assigned to the 2.3.4.4b clade. A variant and SNP analysis found mutations consistent with either avian or cattle origin; mainly, the presence of a glutamic acid, instead of a lysine, in position 627 of the PB2 gene supports a non-human source (Supplemental Table 1).^18^

In conclusion, we report the widespread detection of Influenza A H5N1 virus in wastewater from nine U.S. cities during the spring of 2024. Although the exact cause of the signal is currently unknown, lack of clinical burden along with genomic information suggests avian or bovine origin. Given the now widespread presence of the virus in dairy cows, the concerning findings that unpasteurized milk may contain live virus, and that these two recent factors will increase the number of viral interactions with our species, wastewater monitoring should be readily considered as a sentinel surveillance tool that augments and accelerates our detection of evolutionary adaptations of significant concern.

**Supplementary Figure 1:**
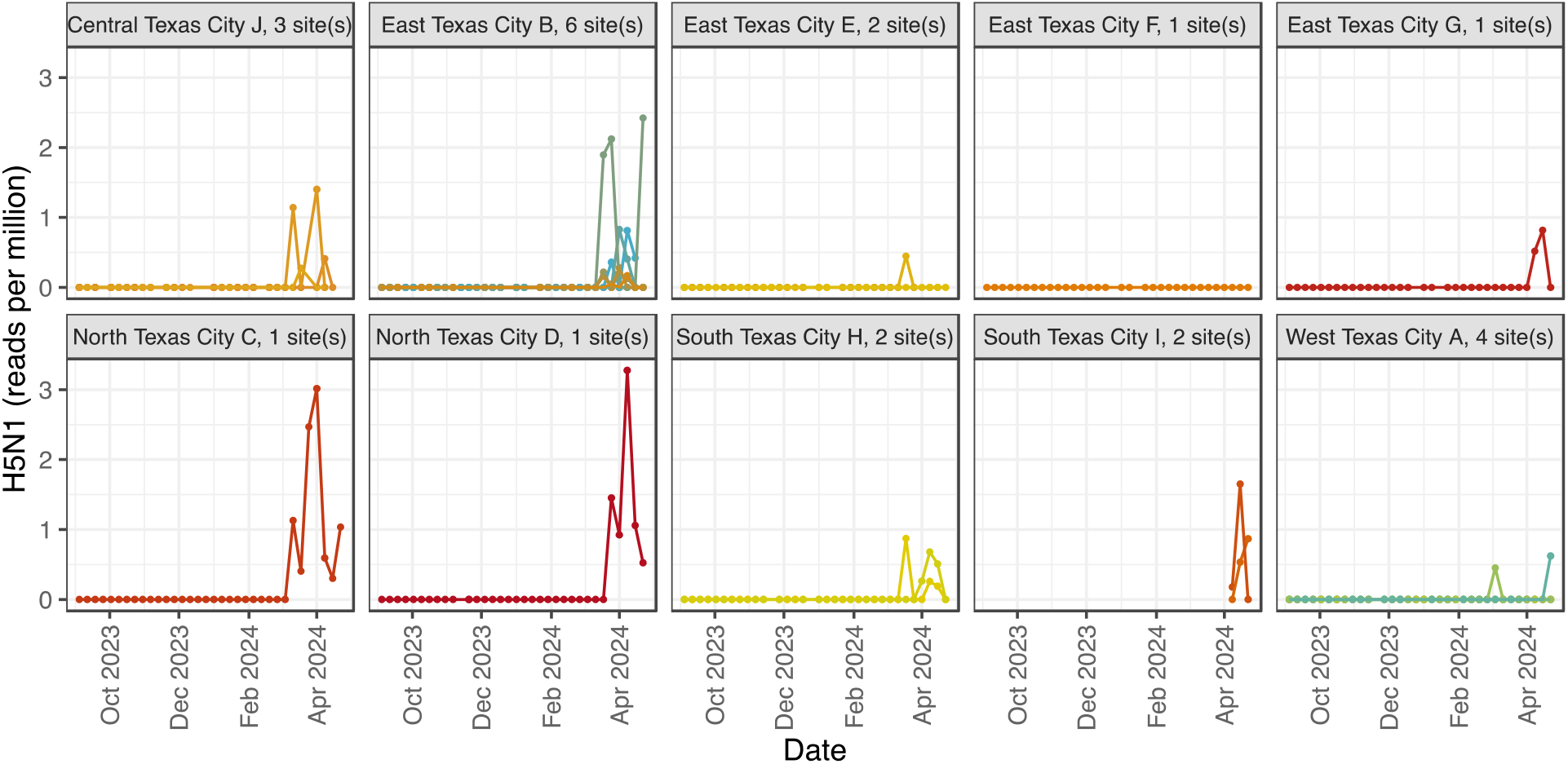
Site-by-site (n=23) detection and quantification of H5N1 sequencing reads from September 1^st^, 2023 to April 25^th^, 2024.

**Supplemental Table 1:**
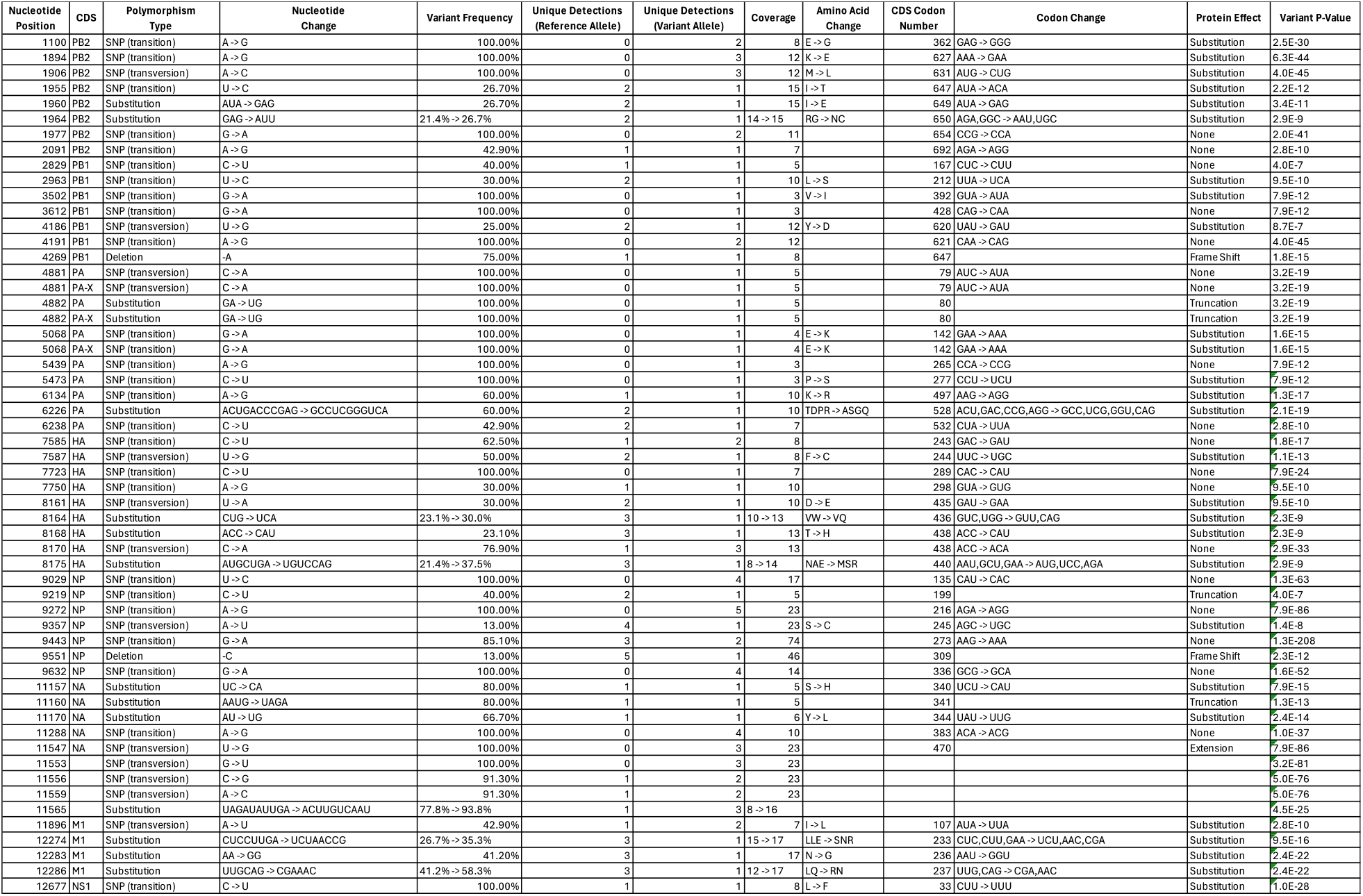
Variant Analysis of Pooled H5N1 Reads Detected in Wastewater. The table presents the results of a variant analysis of H5N1 detected in wastewater. The ‘Nucleotide Position’ refers to the concatenated position in the GISAID EPI_ISL_19027114 genome. ‘Polymorphism Type’ describes the type of genetic variation, with SNP standing for Single Nucleotide Polymorphism. ‘Change’ indicates the alteration in the nucleotide sequence. ‘Variant Frequency’ is the proportion of reads that contain the variant. ‘Unique Detections’ represents the number of times the reference or variant alleles were detected in the read data on different days and/or at different sites. ‘Coverage’ is the number of reads that include the site of the variant. ‘Amino Acid Change’ and ‘CDS Codon Number’ describe the resulting change in the protein sequence and the position of the changed codon in the coding sequence, respectively. ‘Codon Change’ indicates the alteration in protein sequence. ‘Protein Effect’ describes the impact of the variant on the protein, such as a substitution or a frameshift. ‘Variant P-Value’ is an approximate value representing the probability of a sequencing error resulting in observing bases with at least the given sum of qualities. The lower the p-value, the more likely the variation at the given position represents an allele. Abbreviations: CDS, Coding Sequence; SNP, Single Nucleotide Polymorphism; PB2, Polymerase Basic Protein 2; PB1, Polymerase Basic Protein 1; PA, Polymerase Acidic Protein; HA, Hemagglutinin; NP, Nucleoprotein; NA, Neuraminidase; M1, Matrix Protein 1; NS1, Non-structural Protein 1; PA-X, Polymerase Acidic Fusion Protein.

## Methods

### Sample collection and processing

All collection, processing, and sequencing methods have been described in an earlier publication^17^ and were followed precisely. To restate, between 100–500 mL of raw wastewater influent was collected into 500 ml leak-proof prelabeled sample bottles at 23 wastewater treatment plants in 10 Texas cities, the identities of these cities are masked on the request of local public health officials and water utilities. From South Texas City H, sewage from three treatment plants is combined into a single sample before shipment to the sequencing laboratory; all other sites had samples processed individually. Treatment plants were coded upon the request of public health officials. The surface of the sample bottles was decontaminated with 10% bleach and moved to a “clean” zone, where the samples were sealed into biohazard bags in shipping boxes with absorbent pads and ice packs for overnight shipping to the Alkek Center for Metagenomics and Microbiome Research at Baylor College of Medicine, Houston, TX.

Wastewater samples were barcoded upon arrival and stored at 4 °C until processing. First, 50 mL of wastewater was decanted and centrifuged at 3374 x g for 10 min, separating the solid and liquid fractions. The supernatant was then vacuum filtered using an ion-based cellulose filter paper and the virus-containing cellulose filter was placed into a bead-beating tube with lysis buffer. The tube was run on a homogenizer for 1 min at 5 m/s, rested for 1 min, then run on the homogenizer for 1 more minute. Following bead beating the samples were centrifuged at 14– 17×1000 RPM for 2 min. DNA and RNA were extracted using the Qiagen QIAamp VIRAL RNA Mini Kit.

### Library preparation, probe-based virome capture and sequencing

RNA extracts were converted to cDNA using Protoscript II First Strand cDNA Synthesis Kit (New England Biolabs Inc.), NEBNext Ultra II Non-Directional RNA Second Strand Module (New England Biolabs Inc.), and Random Primer 6 (New England Biolabs Inc.). A total of 25 ng of the cDNA and DNA mix was used for library construction using Twist Library Preparation EF 2.0 Kit and Twist Universal Adaptor System (Twist Biosciences). The libraries were pooled, a maximum of 18 samples per pool, at equal mass to a total 1,500 ng per pool. The Twist Comprehensive Viral Research Panel (Twist Biosciences) was used to hybridize the probes at 70 °C for 16 h. The post-capture pool was further PCR amplified for 12 cycles and final libraries were sequenced on Illumina NovaSeq 6000 SP flow cell, to generate 2×150 bp paired-end reads. Following sequencing, raw data files in binary base call (BCL) format were converted into FASTQs and demultiplexed based on the dual-index barcodes using the Illumina ‘bcl2fastq’ software.

### Influenza A read detection and serotype assignment

Reads were processed and analyzed using the publicly available iav_serotype (v0.1.1) software package (https://github.com/mtisza1/influenza_a_serotype). For context, iav_serotype utilizes a database of 207,488 publicly available, complete Influenza A segments from all known serotypes (Influenza_A_segment_sequences database v1.1, available on [Zenodo](https://zenodo.org/records/11123391)). In detail, paired-end reads from wastewater virome sequencing are quality filtered using fastp (v0.23.4) ^19^ with default settings. Reads are aligned to Influenza_A_segment_sequences database v1.1 using minimap2^20^ (v2.28-r1209) and flags “-cx sr --secondary=yes” to allow secondary alignments. Average nucleotide identity (ANI) and alignment fraction (AF) of each alignment are calculated. Then read alignments are assigned to a particular serotype if the best alignment is exclusive to one serotype and (ANI*AF >= 0.9). Else, reads are assigned as “ambiguous”.

### Manual Inspection of Putative H5N1 Sequences

After automated (iav_serotype) detection of H5N1 in wastewater samples was noted by MJT, sequencing reads were shared for independent validation by co-authors BMH and JRC. After sequence inspection confirmed the general findings, parameters for final validation were agreed upon. Using web BLAST^21,22^ against all of NT (final inspection on May 4^th^ and 5^th^), forward and reverse reads from each pair were inspected separately. PCR duplicates were assumed to share origin. Read pairs were marked as “H5N1 specific”, otherwise marked as “ambiguous”. “H5N1 specific” label was used only if neither forward or reverse read alignments had a higher bit score to non-H5N1 genomes and either of the following conditions were met: 1) forward and/or reverse reads aligned with 100% identity and alignment fraction to H5N1genomes and alignments to other serotype were worse by 1 or more bases. 2) forward and/or reverse reads aligned with less than 100% identity to H5N1 and non-H5N1 alignments were worse by 2 or more bases. Of 49 sequencing libraries with automatically assigned reads (per iav_serotype), 46 libraries were manually validated as containing H5N1 specific reads.

### Variant Analysis of Pooled Reads

The variant analysis was conducted on pooled reads, independent of the serotype assignment. Here, raw reads underwent quality trimming to a score of 20 and reads with fewer than 30 base pairs were discarded using BBDuk (v38.84)^23^. Duplicate reads were removed using Dedupe (v38.84). The quality-controlled reads were then pooled and aligned to the GISAID EPI_ISL_19027114 lineage using the Geneious Assembler in Geneious Prime 2024.0.5, with medium sensitivity settings to detect variations of any size.

We excluded variant calls with a Strand Bias P-value less than 0.05 and those resulting from ambiguities in the reference sequence. Strand Bias P-value assesses whether observed strand bias is likely due to chance or systematic effects. Variants with significant strand bias were filtered out to maintain data quality.

### Influenza Hospitalization Data

Weekly statewide and regional hospitalization data was downloaded directly from the Texas Respiratory Illness Interactive Dashboard (https://texas-respiratory-illness-dashboard-txdshsea.hub.arcgis.com/) on May 4^th^, 2024.

## Data Availability

All data produced are available online at
https://zenodo.org/doi/10.5281/zenodo.11175923
and
NCBI SRA BioProject: PRJNA966185

## Acknowledgements

The authors thank the health departments and water utilities of all TEPHI network cities and counties for their support in the contribution of wastewater samples to the project.

## Author Contributions

Conceptualization: M.J.T., B.M.H., M.C.R., S.J.J-C., P.A.P., A.T., A.W.M., J.F.P., E.B.

Methodology: M.J.T., B.M.H., K.P., L.W., M.C.R., S.J.J-C., V.A., A.W.M., J.F.P., E.B., F.W.

Investigation: M.J.T., B.M.H., J.R.C., A.W.M.

Visualization: M.J.T., B.M.H., J.R.C., A.W.M.

Funding acquisition: B.M.H., J.F.P., J.D., E.B., A.W.M., P.A.P.

Project administration: J.D., J.F.P., A.W.M.

Supervision: M.J.T., M.C.R., J.F.P., E.B., A.W.M..

Writing—original draft: M.J.T., B.M.H., J.R.C., A.W.M..

Writing—review and editing: M.J.T., B.M.H., J.R.C., K.P., L.W., J.D., J.C., P.A.P., M.C.R., P.A.P., A.G., A.W.M., J.F.P., J.R., E.B., F.W.

## Data Availability

Read data from virome sequencing of all wastewater samples will be publicly available and deposited in SRA under project accession PRJNA966185.

H5N1 reads by sample are deposited at https://zenodo.org/doi/10.5281/zenodo.11175923.

